# Detection of Novel Acoustic Biomarkers for Parkinson’s Disease through a Machine Learning-Based Composite Spectrogram Analysis

**DOI:** 10.64898/2026.01.01.25343300

**Authors:** Kotaro Tsutsumi, Peter D. Chang, Sanaz Attaripour Isfahani

## Abstract

**Background:** Speech abnormalities are common in Parkinson’s disease (PD). Machine learning (ML) offers potential for objective and scalable speech-based diagnostics. This study introduces an explainable ML pipeline that leverages a novel vowel articulation-based composite input to detect PD and identify phoneme-level biomarkers.

**Methods:** Two publicly available datasets of PD speech recordings were analyzed. Sustained vowel articulations were converted into log-mel spectrograms either individually or as a composite image by vertically concatenating a set of vowels per subject. Processed spectrograms were used to train ML models, with performance assessed using five-fold cross-validation and bootstrapped area under the receiver operating characteristics curve (AUROC). Gradient-weighted Class Activation Mapping (Grad-CAM) was applied to quantify model attention across vowel regions.

**Results:** A total of 150 patients (49.3% PD) were included. Acoustic analysis revealed significant group differences in cepstral peak prominence and harmonics-to-noise ratio, particularly for vowel /u/ (p < 0.05). The ML model achieved an average AUROC of 0.805 using individual vowels and improved to 0.928 with the composite input (p < 0.001). Grad-CAM demonstrated the highest activation for vowel /u/ (p < 0.001), consistent with acoustic findings.

**Conclusion:** The proposed explainable composite spectrogram approach dually enabled high classification performance and identification of a vowel biomarker. Concordance between ML and acoustic analyses highlights the translational potential of explainable ML in PD speech assessment and its ability to reveal underlying pathophysiological insights.

## Introduction

Parkinson’s disease (PD) is a progressive neurodegenerative disorder characterized by the degeneration of dopaminergic neurons in the substantia nigra, resulting in both motor and non-motor symptoms. Cardinal motor features include bradykinesia, resting tremor, rigidity, postural instability, and speech abnormalities, while non-motor symptoms may involve mood disorders, sleep disturbances, and autonomic dysfunction. PD affects millions of individuals worldwide and has a profound impact on functional independence and quality of life. As the global population ages, the prevalence of PD is expected to increase, underscoring the need for improved diagnostic and monitoring tools.^1,2^ Traditionally, PD has been diagnosed and monitored through clinical assessments, such as the Movement Disorder Society Unified Parkinson’s Disease Rating Scale (MDS-UPDRS), which rely on examiner observation and patient-reported symptoms.^3^ While these tools are well-validated and widely used, they are inherently subjective and susceptible to inter-rater variability. This has led to growing interest in developing more objective, quantifiable methods for PD evaluation.

In recent years, advances in machine learning (ML) have enabled the development of tools that can extract meaningful patterns from biological, physiological, and behavioral data to support diagnosis and disease tracking. For instance, studies have utilized multimodal data including proteomics, metabolomics, and genetic data in conjunction with clinical information to predict PD diagnosis and cognitive impairment among people with PD.^4,5^ Furthermore, video-based analysis of facial expression, gait, and finger-tapping, along with sensor-based quantification of bradykinesia have all shown promising outcomes, suggesting that acquisition and assessment of digital biomarkers may allow for non-invasive, real-time monitoring and diagnosis of PD.^6–9^

Among emerging biomarker targets for ML-driven analysis, speech has garnered particular attention. Hypokinetic dysarthria, a hallmark of PD, is characterized by reduced vocal loudness, monotonicity, imprecise consonant articulation, and a breathy or hoarse voice quality.^10,11^ These abnormalities are present in a large proportion of PD patients and may precede overt motor signs by up to 10 years, making them promising candidates for early detection and longitudinal monitoring.^12^ Importantly, speech analysis is non-invasive, cost-effective, and easily scalable with the use of consumer-grade microphones and telehealth infrastructure. A number of recent studies have explored the use of ML to analyze speech in PD. Costantini et al. compared ML approaches to classify PD speech in ON and OFF medication states, highlighting the clinical relevance of acoustic biomarkers.^13^ Pah et al. demonstrated that phoneme features derived from sustained vowels differentiated PD from controls, achieving classification accuracies above 84%.^14^ Other notable contributions, specifically those that explored the utility of inputting speech data in the form of log-mel spectrograms (LMS), include Rahmatallah et al.’s work targeting PD patients and Eguchi et al.’s differentiation of PD and spinocerebellar ataxia using deep neural networks trained on speech data.^15,16^

Despite promising results, several challenges remain. Many existing models are limited by small, proprietary datasets that restrict reproducibility and clinical applicability. Furthermore, few studies have incorporated explainability tools, making it difficult to interpret model decisions and identify clinically meaningful speech biomarkers. There is also limited exploration of how specific phonemes may differentially contribute to disease detection, and few models have leveraged multiple vowel articulations in a unified framework.

To address these gaps, the present study evaluates an explainable ML pipeline for its ability to classify PD based on sustained vowel articulation. We transform vowel samples into LMS and construct a composite input whereby collections of vowel articulations are integrated into a unified framework. This uniquely serves a dual purpose in enabling explainability via visualization of the model’s attention across phonemes while also enhancing classification performance. Furthermore, we compare these results to those from conventional acoustic feature analysis, confirming the clinical utility of our pipeline and shedding light on the pathophysiological basis underlying dysarthria in PD. Through this approach, we aim to both improve autonomous speech-based classification of PD and enhance interpretability in PD speech analysis, bridging the use of such technology closer to real clinical application.

## Methods

### Speech Database

We utilized two publicly available datasets of voice recordings: the NeuroVoz corpus and the Italian Parkinson’s Voice and Speech (ItalianPVS) dataset.^17,18^ Both datasets contain sustained phonation and structured reading tasks, allowing for linguistically uniform and articulatorily focused comparisons between individuals with and without Parkinson’s disease (PD). For the purposes of this study, only sustained vowel phonation recordings (/a/, /e/, /i/, /o/, /u/) were extracted from each dataset to develop a focused classification pipeline targeting Parkinsonian speech.

The NeuroVoz corpus comprises 2,903 speech recordings collected from 108 native Castilian Spanish speakers, including 53 individuals diagnosed with PD and 55 healthy controls (HC). Given the unavailability of some sustained phonation recordings, a total of 101 patients were enrolled for the current study. All participants were recorded while in their medication "ON" state, 2 to 5 hours after dopaminergic intake. Recordings were conducted in a standardized environment at the Hospital General Universitario Gregorio Marañón digitized at 44.1 kHz sampling rate. The corpus includes several structured speech tasks; however, for this study, we exclusively used the recordings of sustained phonation of the five Spanish vowels. Each vowel was phonated at a comfortable pitch and volume, producing approximately 3 to 4 seconds of steady phonation per recording. Participants were guided through the protocol to ensure consistency and reduce variability. Comprehensive metadata including demographic information, vocal assessments, and clinical characteristics such as Hoehn and Yahr (H&Y) stage and UPDRS-III scores were also recorded for each subject.

The ItalianPVS dataset contains audio recordings from 28 individuals with PD, 22 elderly HC participants with no report of speech or language disorders, and 28 young HC subjects, all of whom are native Italian speakers. We excluded young HC subjects and 1 PD subject to control for age and given the lack of audio files, respectively, leaving a total of 49 patients for the current study. Recordings were conducted under controlled acoustic conditions using a consistent microphone setup at a sampling rate of 16 kHz. For this analysis, only the vowel phonation recordings were used. Each vowel was sustained for several seconds following a deep inhalation to ensure maximal phonatory effort, and the task was performed twice per vowel. The protocol was designed to minimize cognitive demand, and participants were instructed to sustain each vowel until their breath was exhausted for the first trial, and for 5 seconds during the second. All participants were receiving antiparkinsonian therapy at the time of recording.

### Audio Preprocessing

All audio files were initially resampled to a uniform sampling rate of 16 kHz and truncated to 2-second segments to ensure consistent analysis duration. Audio preprocessing was performed using the Parselmouth Python library, which provides a Python interface to Praat acoustic analysis software.^19,20^

### Acoustic Feature Extraction

Formant frequencies were estimated using Praat’s Burg algorithm. We specifically focused on the first two formants (F1 and F2) as they are the most robust and clinically relevant for vowel articulation assessment. Formant values were measured at the temporal midpoint of each sustained vowel to ensure stable phonation and minimize transient effects.

Voice quality was assessed through three key measures. Jitter is represented as cycle-to-cycle variation in frequency, calculated as the average absolute difference between consecutive periods divided by the average period, expressed as a percentage. This measure reflects short-term instability in pitch. Shimmer is represented as cycle-to-cycle variation in amplitude, calculated as the average absolute difference between consecutive peak amplitudes divided by the average amplitude, expressed as a percentage. This measure indicates variations in loudness. Harmonics-to-noise ratio (HNR) is the ratio of periodic components to aperiodic components in the voice signal, measured in decibels. HNR provides an index of voice quality, with lower values indicating more noise and potentially pathological voice production. Finally, Cepstral peak prominence (CPP) was extracted as a measure of overall voice quality and vocal fold adduction. CPP represents the prominence of the peak in the cepstrum corresponding to the fundamental frequency and reflects the clarity of harmonic structures of voice, correlating well with perceptual assessments of voice disorder severity.

Furthermore, two vowel space metrics were calculated to assess articulatory precision.^21^ Vowel articulation index (VAI) was calculated as (F1a + F2i) / (F2a + F1i + F1u + F2u), where the subscripts refer to the respective vowels. VAI provides a normalized measure of vowel space utilization, with smaller values indicating reduced articulatory working space. Vowel space area (VSA) represents the triangular area formed by the F1-F2 coordinates of vowels /a/, /i/, and /u/ in the acoustic vowel space, calculated as ABS[F1i(F2a – F2u) + F1a(F2u – F2i) + F1u(F2i – F2a)]/2. VSA represents the overall size of the articulatory working space, with reduced values indicating centralization of vowel production. Of note, all acoustic features were extracted and analyzed with stratification per sex given variability in characteristics intrinsic to subjects’ sex.

### Data Preprocessing and Augmentation for Deep Learning Analysis

All audio files were resampled to a uniform sampling rate of 16 kHz. To standardize input dimensions across samples, we segmented the audio files into 2 second clips. Each audio segment was zero-padded or truncated to ensure a consistent duration of 2 seconds. To enrich the training set, we employed data augmentation strategies including time-shifting (max ±20% of signal duration) and random pitch shifting within ±7 semitones. 7 semitones were selected as range for pitch shifting given this is the typical vocal semitone difference between male and female.

### Log-Mel Spectrogram Transformation

All resampled and preprocessed audio clips were first converted into spectrograms with short-time Fourier transform and subsequently into LMS using 128 mel filter banks. In a mel spectrogram, frequencies are converted into the mel scale, which approximates the nonlinear frequency resolution of human hearing by allocating greater resolution to lower frequencies. Furthermore, a logarithmic transformation is applied to represent amplitude as decibel to mimic the logarithmic sensitivity of the human auditory system, finally yielding LMS. This perceptually weighted representation is particularly well-suited for use with image-based neural network architectures such as convolutional neural networks (CNNs).

### Composite Input Format for Explainability Analysis

To enable explainability analysis and explore its impact on model performance, we implemented a composite input format wherein LMS from each of the five sustained vowels were vertically stacked along the frequency axis to form a single 2D input image. This stacked representation allowed for simultaneous analysis of all vowel articulations. The resulting composite spectrogram was then resized to 224 × 224 pixels and inputted into CNN. Training and testing were conducted using the same fold-wise training-testing dataset splits and evaluation metrics utilized in the non-stacked analysis, as described below.

### Model Training and Evaluation

We utilized the ResNet50 model pretrained on the ImageNet-1K dataset for the binary classification of PD versus HC speech samples. ResNet50 is a CNN originally proposed by He et al., comprising 50 layers including an initial convolution and max pooling layer, followed by 16 residual blocks grouped into four stages, and ending with a global average pooling layer and a fully connected classification head.^22^ The output layer was modified to perform binary classification, with a softmax activation function used to generate class probabilities corresponding to PD or HC. Cross-entropy loss was used as the objective function during training.

Training and testing were conducted using five-fold cross-validation with patient-level stratification, ensuring each subject appeared in either training or testing partitions but not both. The same training-testing dataset splits per each fold were utilized at a ratio of 80:20 across different evaluation methods. The models were optimized using the Adam optimizer with a learning rate of 0.00005, cross-entropy as loss function, and a batch size of 8. The model was trained for a total of 20 epochs (2940-2980 iterations for non-stacked input; 280-300 iterations for composite input). Model performance was assessed using area under the receiver operating characteristic curve (AUROC). To establish statistical significance and confidence intervals, bootstrap resampling with 1000 iterations was performed for each fold. Individual p-values were calculated per each fold. These five independent p-values were then combined using Fisher’s method, yielding an overall p-value that accounts for evidence across all folds.^23^

### Gradient-weighted Class Activation Mapping Implementation

Gradient-weighted Class Activation Mapping (Grad-CAM) was applied to visualize and quantify the model’s attention across different vowel regions during classification decisions.^24^ Grad-CAM identifies regions in the input spectrogram that contribute most significantly to the model’s classification by computing the gradient of the class score with respect to the feature maps of the last convolutional layer. The target layer for gradient extraction was the final convolutional layer of ResNet50, which captures high-level semantic features while maintaining sufficient spatial resolution for localization.

The composite spectrogram was divided into five equal segments along the frequency axis, each corresponding to one vowel (/a/, /e/, /i/, /o/, /u/). For each segment, we calculated the number of pixels exceeding a predetermined activation threshold, set at 0.8, per each vowel per sample. This was aggregated across all samples to identify group-level differences in vowel-specific model attention patterns, providing insights into potential acoustic biomarkers differentiating PD from HC speech. The Kruskal–Wallis test was employed to statistically assess whether the distribution of Grad-CAM activations differed significantly across vowel categories.

### Statistical Analysis

Univariate analysis was performed using two-tailed Welch’s t-tests to compare acoustic features between PD and HC groups. Given the multiple comparisons performed, Bonferroni correction was applied to control for family-wise error rate. Individual p-values were adjusted accordingly per each variable, and a significance level of 0.05 was used for all tests. All preprocessing, model training, and statistical analyses were implemented in Python (version 3.12.11).

## Results

In total, 150 participants were included in the study, with a mean age of 67.1 ± 9.87 years, and 44% were female (**Table 1**). Among them, 74 participants had a confirmed clinical diagnosis of PD. Within the NeuroVoz cohort, PD participants were significantly older than HC (p = 0.002), while no significant sex differences were observed. In the ItalianPVS dataset, there were no significant differences in age and sex distribution between groups. When pooled, PD participants were significantly older than HC (p = 0.005), with similar sex distribution.

**Table 1.** Demographic Information.

| Dataset | Variables | Total | HC | PD | p-value |
| --- | --- | --- | --- | --- | --- |
| NeuroVoz | Sample size | 101 | 54 | 47 |  |
|  | Age (mean, SD) | 67.1±10.9 | 64±10.4 | 70.6±10.6 | 0.002 |
|  | Sex (Female, %) | 45 (45) | 26 (48) | 19 (40) | 0.436 |
|  | UPDRS-III (mean, SD) |  |  | 17.2±11.8 |  |
|  | H&Y (mean, SD) |  |  | 2.2±0.72 |  |
| ItalianPVS | Sample size | 49 | 22 | 27 |  |
|  | Age (mean, SD) | 67.1±7.37 | 67.1±5.16 | 67.1±8.87 | 0.993 |
|  | Sex (Female; %) | 21 (43) | 12 (55) | 9 (33) | 0.136 |
| Combined | Sample size | 150 | 76 | 74 |  |
|  | Age (mean, SD) | 67.1±9.87 | 64.9±9.21 | 69.3±10.1 | 0.005 |
|  | Sex (Female, %) | 66 (44) | 38 (50) | 28 (38) | 0.134 |

HC: healthy control, H&Y: Hoehn and Yahr, ItalianPVS: Italian Parkinson's Voice and Speech, PD: Parkinson's disease, SD: standard deviation, UPDRS: Unified Parkinson's Disease Rating Scale.

Acoustic analyses comparing vowel features between PD and HC groups were conducted in sex-stratified fashion. Among male participants (**Table 2**), for vowel /a/, F1 was significantly reduced in PD compared to HC (p = 0.032). For vowel /e/, F2 was lower in PD (p = 0.022). Additionally, HNR for vowel /i/ was elevated in PD (p = 0.032). For vowel /u/, both CPP and HNR were significantly higher among PD males (p < 0.001; p = 0.003). In female participants (**Table 3**), CPP for vowel /i/ was greater in PD than HC (p < 0.001). Similar to males, for vowel /u/, both CPP and HNR were elevated in PD (p < 0.001; p = 0.026). No additional acoustic features—including jitter, shimmer, VAI, or VSA—retained significance following correction for multiple-comparison.

**Table 2.**
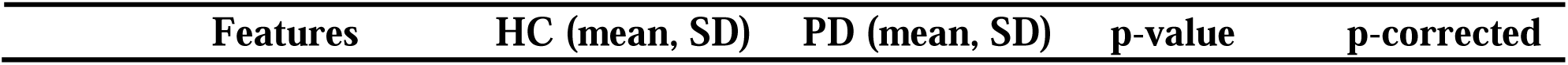
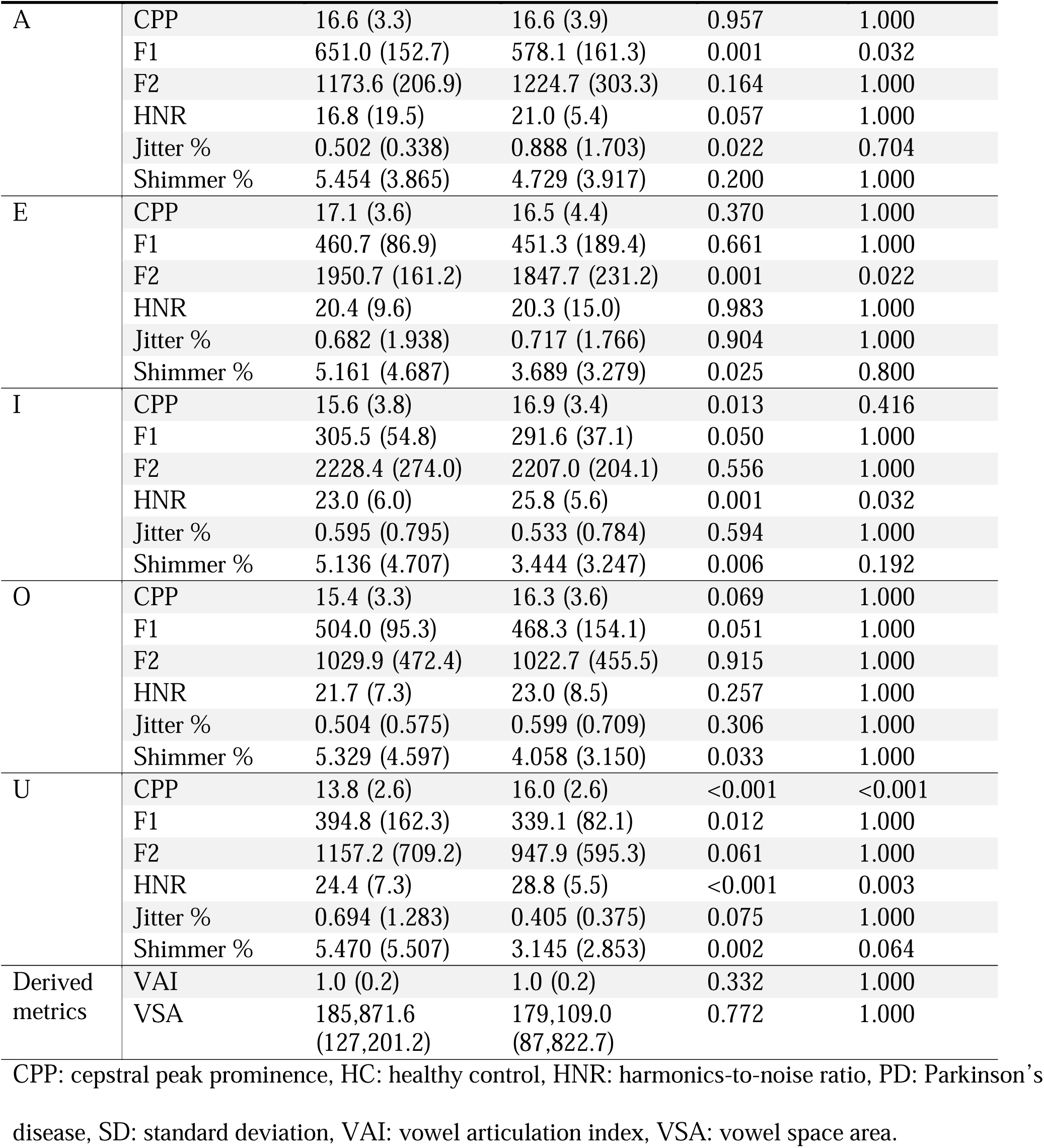
Acoustic Analysis of Vowel Sounds from Male Participants.

| Features | HC (mean, SD) | PD (mean, SD) | p-value | p-corrected |
| --- | --- | --- | --- | --- |

|  |  |  |  |  |  |
| --- | --- | --- | --- | --- | --- |
| A | CPP | 16.6 (3.3) | 16.6 (3.9) | 0.957 | 1.000 |
|  | F1 | 651.0 (152.7) | 578.1 (161.3) | 0.001 | 0.032 |
|  | F2 | 1173.6 (206.9) | 1224.7 (303.3) | 0.164 | 1.000 |
|  | HNR | 16.8 (19.5) | 21.0 (5.4) | 0.057 | 1.000 |
|  | Jitter % | 0.502 (0.338) | 0.888 (1.703) | 0.022 | 0.704 |
|  | Shimmer % | 5.454 (3.865) | 4.729 (3.917) | 0.200 | 1.000 |
| E | CPP | 17.1 (3.6) | 16.5 (4.4) | 0.370 | 1.000 |
|  | F1 | 460.7 (86.9) | 451.3 (189.4) | 0.661 | 1.000 |
|  | F2 | 1950.7 (161.2) | 1847.7 (231.2) | 0.001 | 0.022 |
|  | HNR | 20.4 (9.6) | 20.3 (15.0) | 0.983 | 1.000 |
|  | Jitter % | 0.682 (1.938) | 0.717 (1.766) | 0.904 | 1.000 |
|  | Shimmer % | 5.161 (4.687) | 3.689 (3.279) | 0.025 | 0.800 |
| I | CPP | 15.6 (3.8) | 16.9 (3.4) | 0.013 | 0.416 |
|  | F1 | 305.5 (54.8) | 291.6 (37.1) | 0.050 | 1.000 |
|  | F2 | 2228.4 (274.0) | 2207.0 (204.1) | 0.556 | 1.000 |
|  | HNR | 23.0 (6.0) | 25.8 (5.6) | 0.001 | 0.032 |
|  | Jitter % | 0.595 (0.795) | 0.533 (0.784) | 0.594 | 1.000 |
|  | Shimmer % | 5.136 (4.707) | 3.444 (3.247) | 0.006 | 0.192 |
| O | CPP | 15.4 (3.3) | 16.3 (3.6) | 0.069 | 1.000 |
|  | F1 | 504.0 (95.3) | 468.3 (154.1) | 0.051 | 1.000 |
|  | F2 | 1029.9 (472.4) | 1022.7 (455.5) | 0.915 | 1.000 |
|  | HNR | 21.7 (7.3) | 23.0 (8.5) | 0.257 | 1.000 |
|  | Jitter % | 0.504 (0.575) | 0.599 (0.709) | 0.306 | 1.000 |
|  | Shimmer % | 5.329 (4.597) | 4.058 (3.150) | 0.033 | 1.000 |
| U | CPP | 13.8 (2.6) | 16.0 (2.6) | <0.001 | <0.001 |
|  | F1 | 394.8 (162.3) | 339.1 (82.1) | 0.012 | 1.000 |
|  | F2 | 1157.2 (709.2) | 947.9 (595.3) | 0.061 | 1.000 |
|  | HNR | 24.4 (7.3) | 28.8 (5.5) | <0.001 | 0.003 |
|  | Jitter % | 0.694 (1.283) | 0.405 (0.375) | 0.075 | 1.000 |
|  | Shimmer % | 5.470 (5.507) | 3.145 (2.853) | 0.002 | 0.064 |
| Derived metrics | VAI | 1.0 (0.2) | 1.0 (0.2) | 0.332 | 1.000 |
|  | VSA | 185,871.6 (127,201.2) | 179,109.0 (87,822.7) | 0.772 | 1.000 |

CPP: cepstral peak prominence, HC: healthy control, HNR: harmonics-to-noise ratio, PD: Parkinson's
disease, SD: standard deviation, VAI: vowel articulation index, VSA: vowel space area.

**Table 3.**
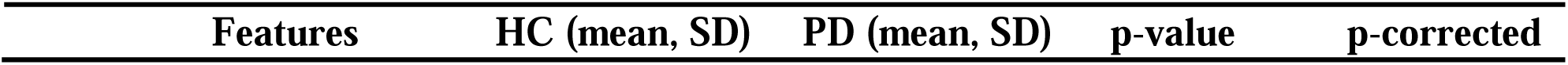
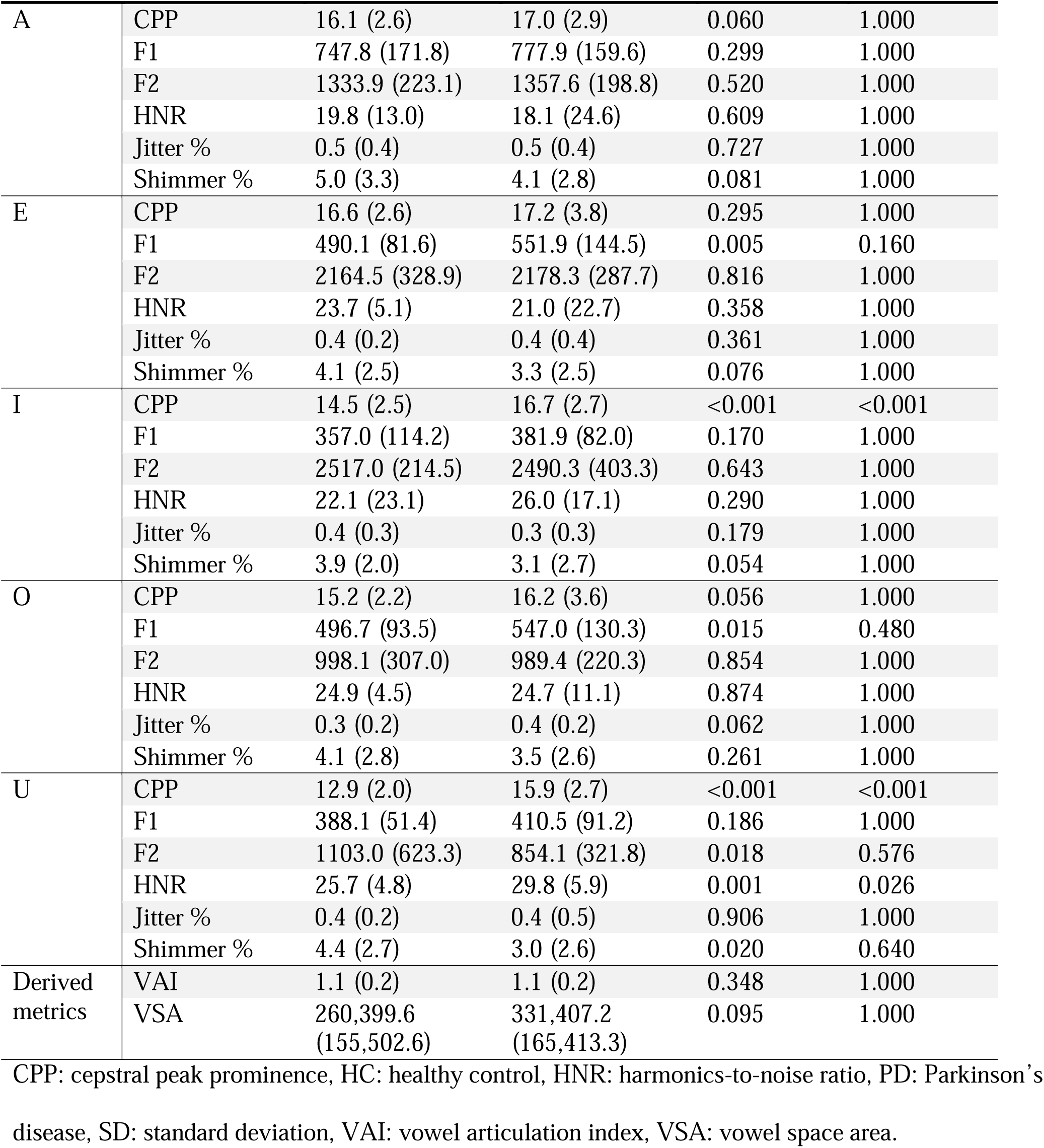
Acoustic Analysis of Vowel Sounds from Female Participants.

| Features | HC (mean, SD) | PD (mean, SD) | p-value | p-corrected |
| --- | --- | --- | --- | --- |

|  |  |  |  |  |  |
| --- | --- | --- | --- | --- | --- |
| A | CPP | 16.1 (2.6) | 17.0 (2.9) | 0.060 | 1.000 |
|  | F1 | 747.8 (171.8) | 777.9 (159.6) | 0.299 | 1.000 |
|  | F2 | 1333.9 (223.1) | 1357.6 (198.8) | 0.520 | 1.000 |
|  | HNR | 19.8 (13.0) | 18.1 (24.6) | 0.609 | 1.000 |
|  | Jitter % | 0.5 (0.4) | 0.5 (0.4) | 0.727 | 1.000 |
|  | Shimmer % | 5.0 (3.3) | 4.1 (2.8) | 0.081 | 1.000 |
| E | CPP | 16.6 (2.6) | 17.2 (3.8) | 0.295 | 1.000 |
|  | F1 | 490.1 (81.6) | 551.9 (144.5) | 0.005 | 0.160 |
|  | F2 | 2164.5 (328.9) | 2178.3 (287.7) | 0.816 | 1.000 |
|  | HNR | 23.7 (5.1) | 21.0 (22.7) | 0.358 | 1.000 |
|  | Jitter % | 0.4 (0.2) | 0.4 (0.4) | 0.361 | 1.000 |
|  | Shimmer % | 4.1 (2.5) | 3.3 (2.5) | 0.076 | 1.000 |
| I | CPP | 14.5 (2.5) | 16.7 (2.7) | <0.001 | <0.001 |
|  | F1 | 357.0 (114.2) | 381.9 (82.0) | 0.170 | 1.000 |
|  | F2 | 2517.0 (214.5) | 2490.3 (403.3) | 0.643 | 1.000 |
|  | HNR | 22.1 (23.1) | 26.0 (17.1) | 0.290 | 1.000 |
|  | Jitter % | 0.4 (0.3) | 0.3 (0.3) | 0.179 | 1.000 |
|  | Shimmer % | 3.9 (2.0) | 3.1 (2.7) | 0.054 | 1.000 |
| O | CPP | 15.2 (2.2) | 16.2 (3.6) | 0.056 | 1.000 |
|  | F1 | 496.7 (93.5) | 547.0 (130.3) | 0.015 | 0.480 |
|  | F2 | 998.1 (307.0) | 989.4 (220.3) | 0.854 | 1.000 |
|  | HNR | 24.9 (4.5) | 24.7 (11.1) | 0.874 | 1.000 |
|  | Jitter % | 0.3 (0.2) | 0.4 (0.2) | 0.062 | 1.000 |
|  | Shimmer % | 4.1 (2.8) | 3.5 (2.6) | 0.261 | 1.000 |
| U | CPP | 12.9 (2.0) | 15.9 (2.7) | <0.001 | <0.001 |
|  | F1 | 388.1 (51.4) | 410.5 (91.2) | 0.186 | 1.000 |
|  | F2 | 1103.0 (623.3) | 854.1 (321.8) | 0.018 | 0.576 |
|  | HNR | 25.7 (4.8) | 29.8 (5.9) | 0.001 | 0.026 |
|  | Jitter % | 0.4 (0.2) | 0.4 (0.5) | 0.906 | 1.000 |
|  | Shimmer % | 4.4 (2.7) | 3.0 (2.6) | 0.020 | 0.640 |
| Derived metrics | VAI | 1.1 (0.2) | 1.1 (0.2) | 0.348 | 1.000 |
|  | VSA | 260,399.6 (155,502.6) | 331,407.2 (165,413.3) | 0.095 | 1.000 |

CPP: cepstral peak prominence, HC: healthy control, HNR: harmonics-to-noise ratio, PD: Parkinson's
disease, SD: standard deviation, VAI: vowel articulation index, VSA: vowel space area.

An overview of our entire PD classification pipeline is represented in **Figure 1**. Model evaluation was performed using five-fold cross-validation with bootstrapping per each fold (**Table 4**). When trained on individual LMS inputs, the ML model achieved an average AUROC of 0.805. Incorporating the composite stacked input, whereby LMS images from all five vowels were vertically concatenated, yielded an average AUROC of 0.928, a statistically significant improvement in classification performance (p < 0.001). Averaged ROC curves using both methods are depicted in **Figure 2**.

**Figure 1.**
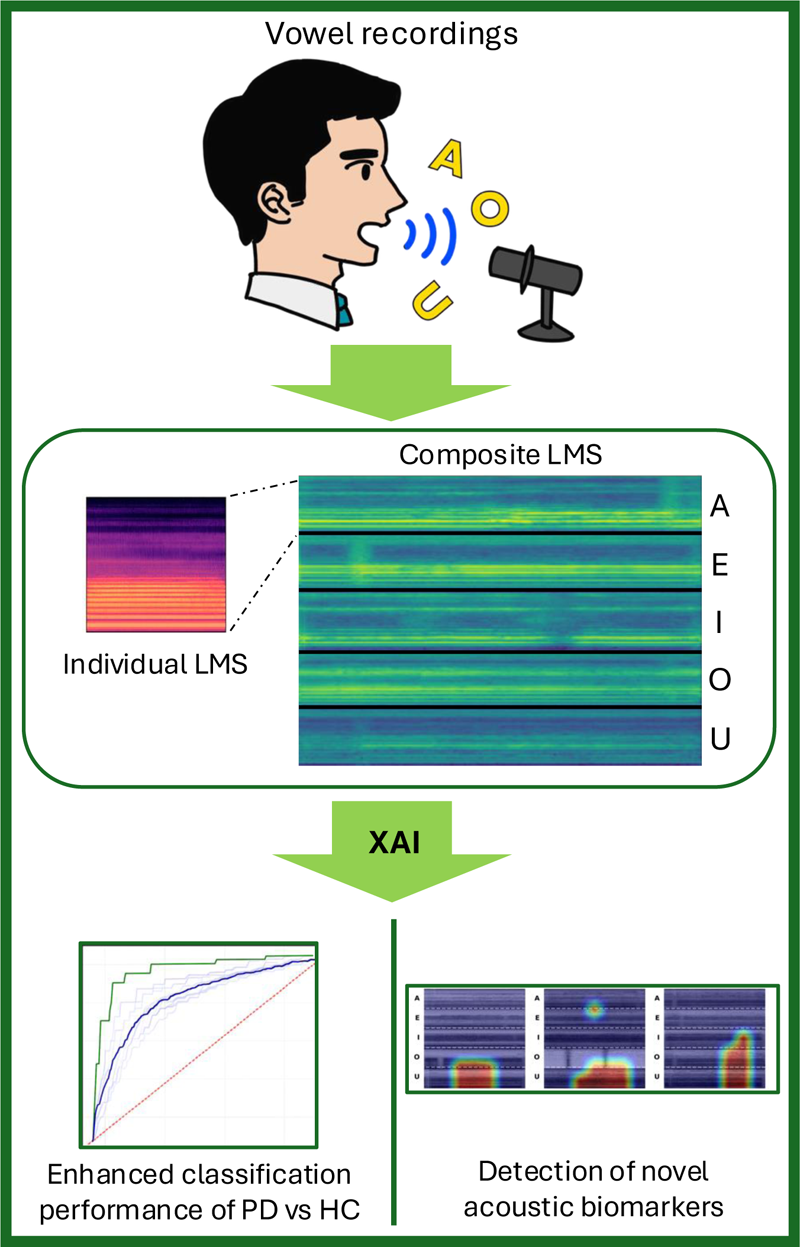
Overview of the PD Classification Pipeline. Vowel recordings are first transformed into LMS. Individual LMS are concatenated vertically to form a composite input. Individual LMS and composite LMS are separately input into ML for training and testing for comparison of performance in classifying PD vs HC samples. XAI is applied for visualization of the model’s attention across phonemes. We show the utility of composite LMS for enhancing classification performance and enabling detection of novel acoustic biomarkers. HC: healthy control, LMS: log-mel spectrogram, ML: machine learning, PD: Parkinson’s disease, XAI: explainable artificial intelligence.

**Figure 2.**
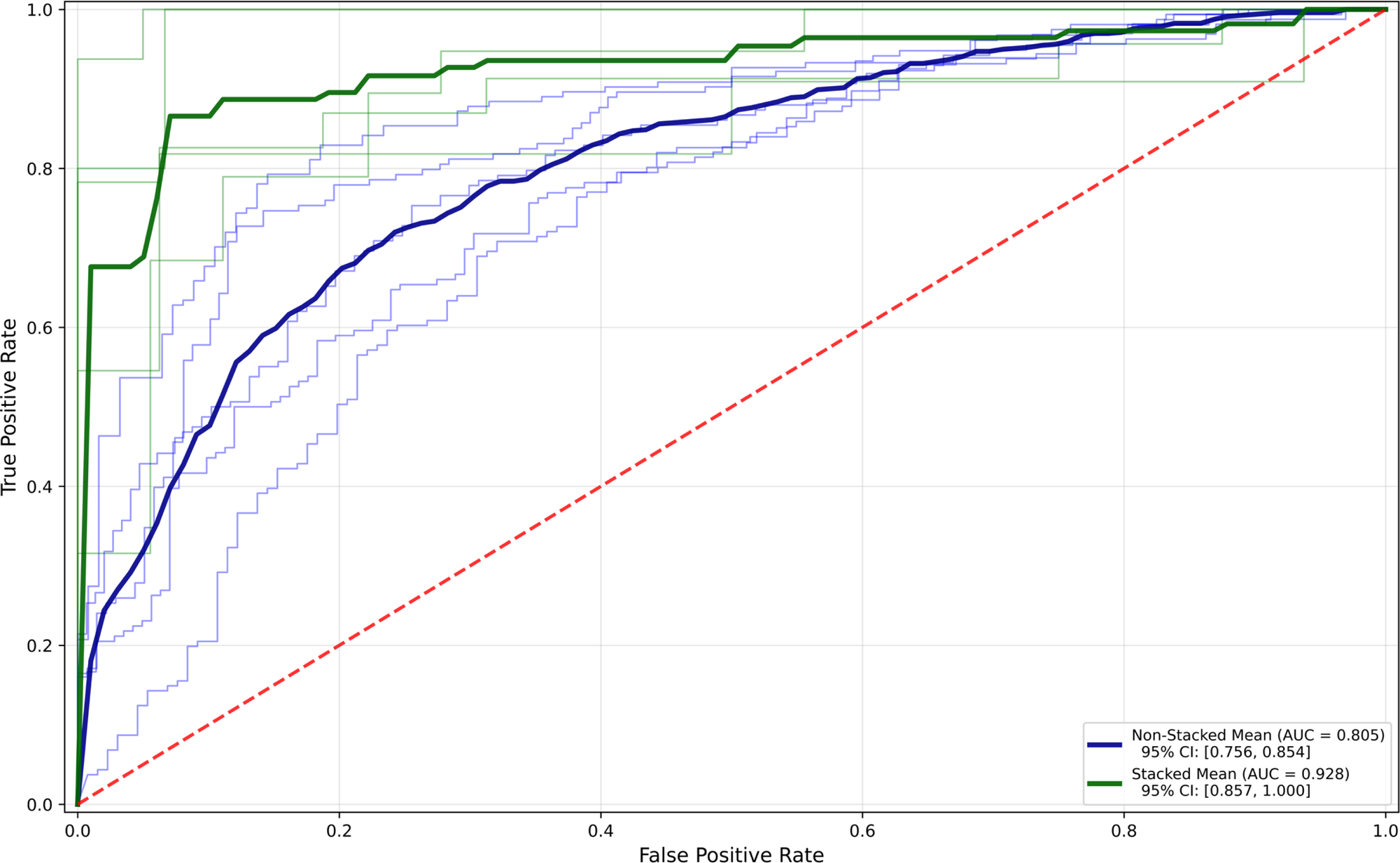
Average ROC Curves for PD Classification. The blue line represents the ROC curve averaged across a five-fold cross validation when using the non-stacked LMS as inputs. The green line represents the ROC curve averaged across a five-fold cross validation when using the stacked composite images as inputs. AUC: area under the curve, CI: confidence interval, LMS: log-mel spectrogram, PD: Parkinson’s disease, ROC: receiver operating characteristic.

**Table 4.**
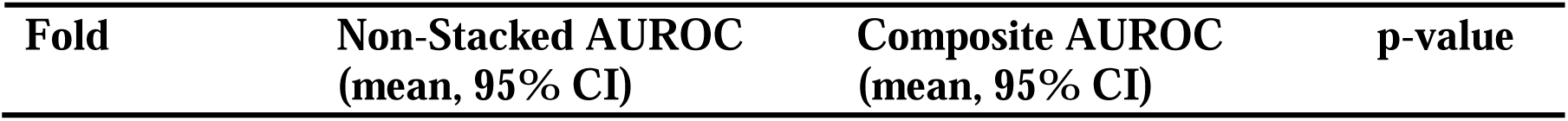
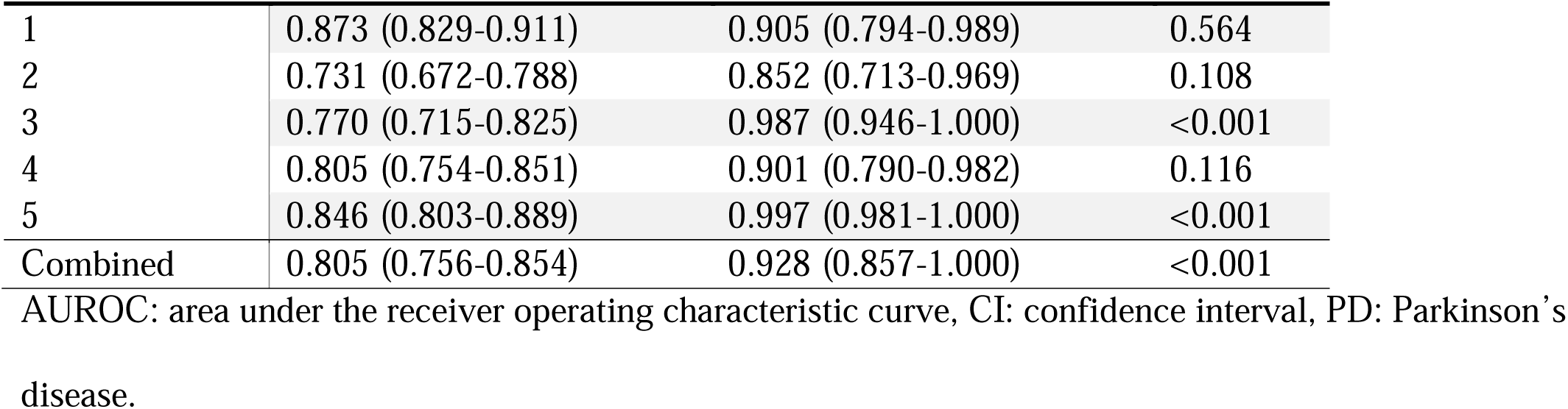
Classification Performance for Detecting PD.

Grad-CAM was employed to visualize the model’s spatial attention across the composite spectrograms. The heatmaps were partitioned into five equal frequency-axis segments corresponding to each vowel. The proportion of pixels exceeding an activation threshold of 0.8 was quantified per vowel and aggregated across samples. Representative images of heatmaps belonging to top 10 samples with highest predictive probabilities per each cohort are displayed in **Figure 3**. Among the HC, PD, and combined cohorts, the Kruskal–Wallis test demonstrated significant non-uniformity in attention distribution (p < 0.001), indicating that specific vowel regions may have contributed more prominently to model decision-making (**Table 5**; **Figure 4**). Among all cohorts, /u/ exhibited the greatest mean activation.

**Figure 3.**
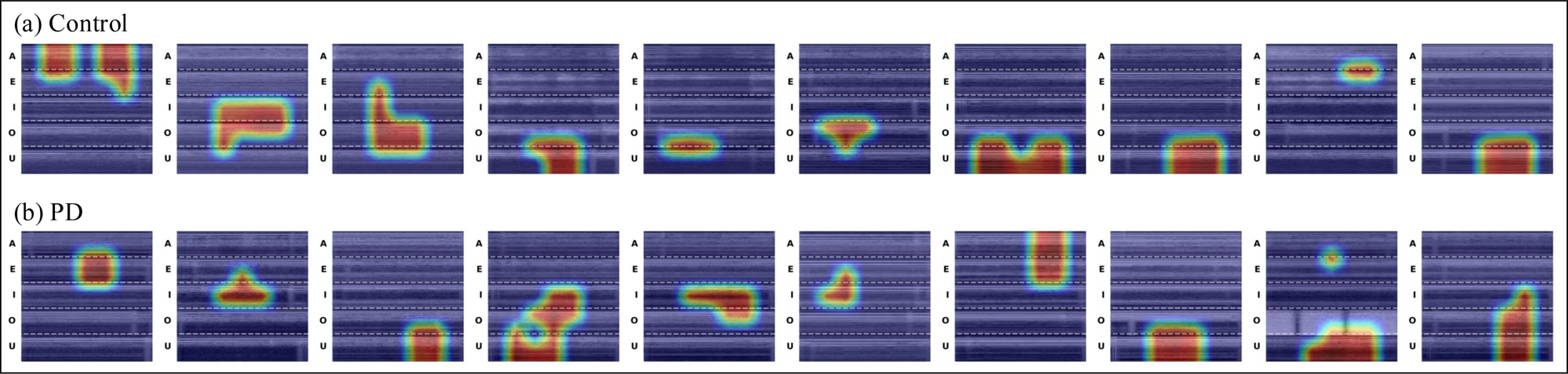
Representative Grad-CAM Activation Maps. Individual LMS images were vertically stacked and concatenated to yield a single composite input that integrated information from all vowel sounds. Grad-CAM was applied to visualize the model’s spatial attention across these composite spectrograms, specifically regions exceeding an activation threshold of 0.8. (a) and (b) respectively represents control and PD samples with highest classification probabilities among each testing cohort. AUC: area under the curve, CI: confidence interval, Grad-CAM: Gradient-weighted Class Activation Mapping, LMS: log-mel spectrogram, PD: Parkinson’s disease, ROC: receiver operating characteristic.

**Figure 4.**
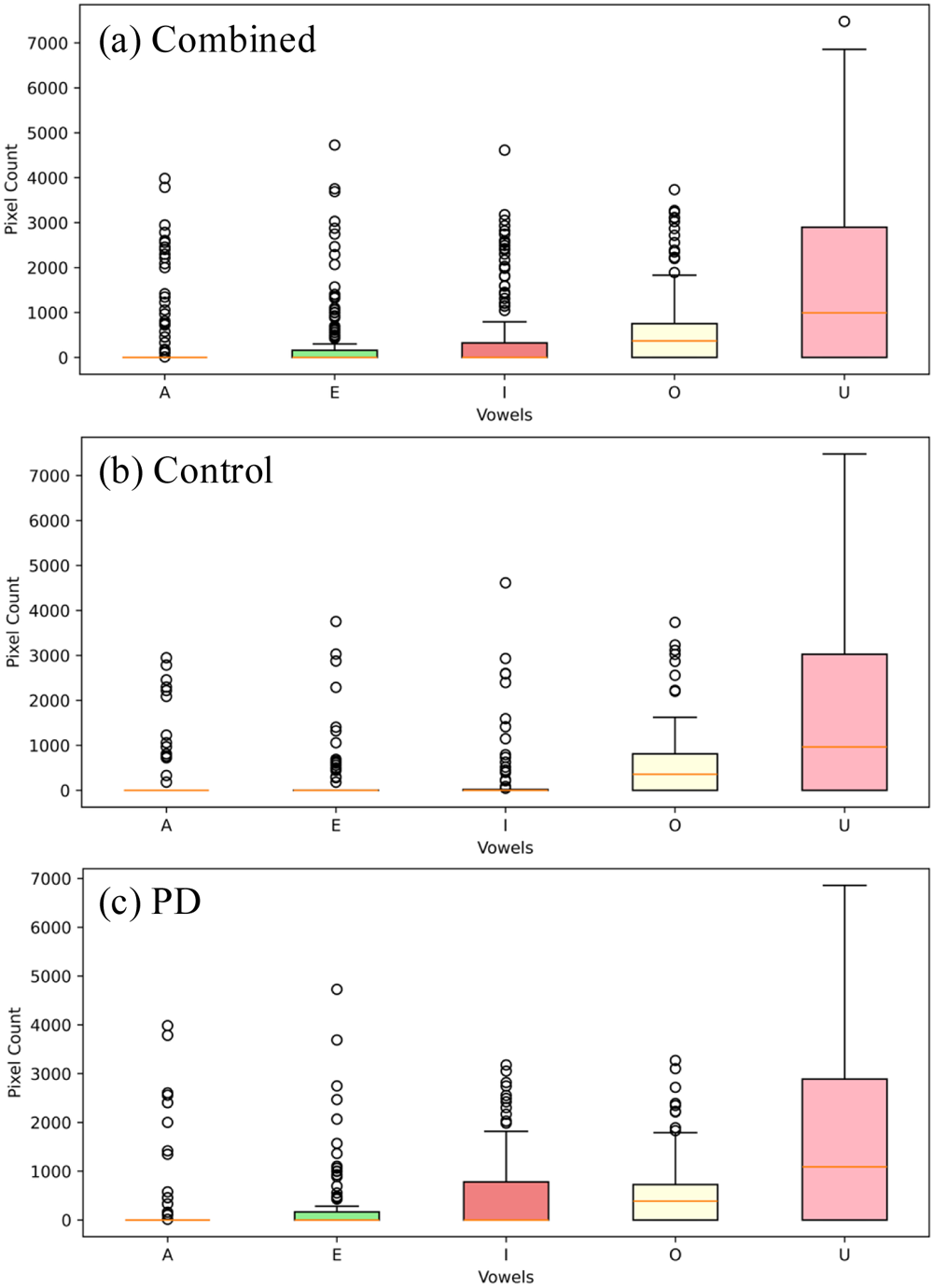
Box Plot of Summative Grad-CAM Activations Across Vowel Sounds. The number of pixels exceeding Grad-CAM’s activation threshold of 0.8 was quantified and summed across all cross-validation folds per vowel sound. Results are represented through a box plot with their quartiles. (a) Combined cohort. (b) Control cohort. (c) PD cohort. Grad-CAM: Gradient-weighted Class Activation Mapping, PD: Parkinson’s disease.

**Table 5.** Grad-CAM Activations per Vowel in Pixels (Activation Threshold = 0.8).

| Cohort | A (mean, SD) | E (mean, SD) | I (mean, SD) | O (mean, SD) | U (mean, SD) | p-value |
| --- | --- | --- | --- | --- | --- | --- |
| HC | 282 (708) | 284 (726) | 316 (826) | 676 (894) | 1641 (1889) | <0.001 |
| PD | 301 (834) | 346 (832) | 541 (915) | 623 (807) | 1680 (1904) | <0.001 |
| Combined | 292 (775) | 317 (782) | 434 (879) | 648 (847) | 1661 (1891) | <0.001 |

Grad-CAM: Gradient-weighted Class Activation Mapping, HC: healthy control, PD: Parkinson's disease, SD: standard deviation.

## Discussion

This study demonstrates the utility of vowel articulation-based ML analysis for differentiating PD from HC while simultaneously uncovering acoustic biomarkers through explainability-driven visualization. We transformed sustained vowel phonations into LMS and uniquely employed a composite input representation that integrates all vowel articulations into a unified framework. Utilizing this composite format not only enhanced classification performance but also enabled explainability through Grad-CAM visualizations, facilitating direct quantification of model attention to specific vowel sounds. The approach highlights how our interpretable pipeline can serve as both a diagnostic and a translational tool, ultimately bridging the gap between informatics and real-world clinical application.

LMS offers an image-based representation of audio features, encompassing time, frequency, and intensity. Rather than compressing these into a discrete and fixed set of features, such as mel-frequency cepstrum coefficients, jitter, shimmer, and formants, LMS’s time-frequency structure allows for preservation of relevant information such as local dependencies and alterations in harmonics. This may enable ML to extract temporal and spectral patterns reflective of dysarthric features that may otherwise be diluted when represented through averaged acoustic parameters.^25^ Importantly, in our study, using the composite spectrogram input—constructed by vertically stacking LMS from the five sustained vowels—yielded a statistically significant improvement in AUROC compared to when individual LMSs were used. This approach integrates phonemic diversity into a single view, perhaps allowing the network to learn inter-vowel relationships and global articulation trends while maintaining localized vowel-specific information. The unified format also supports explainability given that all vowel regions coexist within a shared activation map, enabling quantitative localization of model focus across vowel types. Thus, our original design provides dual benefits, first in performance augmentation, and second in enabling interpretability by revealing which vowel sound contributes most significantly to classification.

Explainability analysis using Grad-CAM revealed that the vowel /u/ consistently elicited the greatest model activation across PD and HC samples. This finding suggests that /u/ may serve as a particularly sensitive acoustic biomarker for distinguishing PD-related dysarthria. The vowel /u/ is characterized with high and backward tongue positioning while also requiring precise control of lip rounding.^26^ These movements may have more demanding involvement of orofacial muscles, making them susceptible to rigidity and reduced motor flexibility in PD. Furthermore, studies have hypothesized that involvement of posterior tongue root may be implicated in early speech and swallowing abnormalities seen in PD.^27,28^ Since vowel /u/ requires posterior tongue elevation, a similar pathophysiological mechanism may be facilitating dysphagia and the selective articulatory impairment of this vowel sound. Our finding is consistent with a previous study that conducted formant-based vowel articulation analysis and found /u/ to be the most reliably impacted vowel sound among others.^10^

Notably, conventional acoustic feature analysis corroborated this model-driven finding. Vowel sound /u/ exhibited the most consistent number of significantly different acoustic features in both sexes, notably with significant elevation of both CPP and HNR. This congruence between explainability analysis and feature-level quantification underscores the reliability of Grad-CAM based strategy as a potential biomarker discovery tool. Traditionally, CPP and HNR are reported to be reduced in pathologic speech, reflecting reduced phonatory stability, decreased harmonic energy, increased noise components, and overall breathiness due to glottal irregularity, though longitudinal studies involving PD patients are lacking.^29^ Our cohort exhibited an inverse trend, with PD speakers demonstrating higher CPP and HNR, particularly for /u/. While PD patients typically show reduced vocal intensity overall, the increased vocal fold tension and rigidity may paradoxically promote more stability of harmonics and periodicity, albeit being strained. This may contrast to running speech, where PD-related movement deficits and fatigue contribute to irregular vocal onset and reduced periodicity. Thus, our findings from vowel articulation tasks may be uniquely influenced by rigidity and phonatory effort, yielding acoustic measures that deviate from expectations established in spontaneous or connected speech. Higher quantitative acoustic measures may represent altered motor control and pathological compensation rather than improved voice quality, and vowel /u/ might be particularly sensitive to detecting such early compensatory changes.

Several limitations should be considered when interpreting the present findings. First, although two publicly available datasets were utilized to enhance sample diversity and reproducibility, the overall dataset size remained modest and there lacked a dedicated external validation dataset. This may have constrained the model’s generalizability. Second, both datasets were collected externally rather than in-house, limiting experimental control over recording protocols, equipment standardization, and participant selection. As a result, the acoustic characteristics and ML-derived features identified here may partly reflect dataset-specific properties. Finally, the ItalianPVS dataset contained limited clinical PD-relevant data such as UPDRS-III scores, H&Y stage, and medication on-off state, preventing detailed correlation between speech-based biomarkers and disease characteristics. Future work leveraging larger, prospectively collected, and clinically annotated datasets is warranted to confirm the reproducibility and translational potential of the identified biomarkers.

## Conclusion

This study illustrates how integrating an image-based explainable ML pipeline with conventional acoustic analysis can advance understanding of PD speech pathophysiology. Our unique composite spectrogram input dually achieved improved classification performance and detection of a phoneme-level biomarker, while alignment between ML-focused vowel sound and statistically significant acoustic features underscored its validity. Our pipeline contributes to both diagnostic automation and pathophysiological insight, bridging the gap between ML analysis and clinically interpretable biomarkers.

## Author Contributions

K.T. contributed to study design, data acquisition, statistical analysis, data interpretation, drafting, and finalizing the manuscript. P.D.C contributed to study design, data interpretation, revising, and finalizing the manuscript. S.A.I. contributed to study design, data interpretation, revising and finalizing the manuscript.

## Author Approval

All authors have seen and approved the manuscript.

## Acknowledgements

None.

## Data Availability Statement

Data from the NeuroVoz corpus and the Italian Parkinson’s Voice and Speech dataset utilized in this study are available from corresponding study groups.

## Code availability

Details regarding our ML framework are described in the Methods section. The codes used in this study are not publicly available but may be available to qualified researchers from the corresponding author upon reasonable request.

## References

1. Dorsey, E. R., and Bloem, B. R. (2018). The Parkinson pandemic—a call to action. JAMA Neurol. 75, 9–10. doi: 10.1001/jamaneurol.2017.3299

2. Bloem, B. R., Okun, M. S., and Klein, C. (2021). Parkinson’s disease. Lancet 397, 2284–2303. doi: 10.1016/S0140-6736(21)00218-X

3. Goetz, C. G., Tilley, B. C., Shaftman, S. R., Stebbins, G. T., Fahn, S., Martinez-Martin, P., et al. (2008). Movement Disorder Society-sponsored revision of the Unified Parkinson’s Disease Rating Scale (MDS-UPDRS): scale presentation and clinimetric testing results. Mov. Disord. 23, 2129–2170. doi: 10.1002/mds.22340

4. Makarious, M. B., Leonard, H. L., Vitale, D., Iwaki, H., Sargent, L., Dadu, A., et al. (2022). Multi-modality machine learning predicting Parkinson’s disease. NPJ Parkinsons Dis. 8:35. doi: 10.1038/s41531-022-00288-w

5. Yang, B., Zhu, Y., Li, K., Wang, F., Liu, B., Zhou, Q., et al. (2024). Machine learning model based on metabolomics and proteomics to predict cognitive impairment in Parkinson’s disease. NPJ Parkinsons Dis. 10:187. doi: 10.1038/s41531-024-00795-y

6. Eguchi, K., Takigawa, I., Shirai, S., Takahashi-Iwata, I., Matsushima, M., Kano, T., et al. (2023). Gait video-based prediction of unified Parkinson’s disease rating scale score: a retrospective study. BMC Neurol. 23:358. doi: 10.1186/s12883-023-03385-2

7. Yang, Y. Y., Ho, M. Y., Tai, C. H., Wu, R. M., Kuo, M. C., and Tseng, Y. J. (2024). FastEval Parkinsonism: an instant deep learning-assisted video-based online system for Parkinsonian motor symptom evaluation. NPJ Digit. Med. 7:31. doi: 10.1038/s41746-024-01022-x

8. Morinan, G., Dushin, Y., Sarapata, G., Rupprechter, S., Peng, Y., Girges, C., et al. (2023). Computer vision quantification of whole-body Parkinsonian bradykinesia using a large multi-site population. NPJ Parkinsons Dis. 9:10. doi: 10.1038/s41531-023-00454-8

9. Hoffman, S. L., Schmiedmayer, P., Gala, A. S., Wilkins, K. B., Parisi, L., Karjagi, S., et al. (2024). Comprehensive real time remote monitoring for Parkinson’s disease using Quantitative DigitoGraphy. NPJ Parkinsons Dis. 10:137. doi: 10.1038/s41531-024-00751-w

10. Rusz, J., Cmejla, R., Tykalova, T., Ruzickova, H., Klempir, J., Majerova, V., et al. (2013). Imprecise vowel articulation as a potential early marker of Parkinson’s disease: effect of speaking task. J. Acoust. Soc. Am. 134, 2171–2181. doi: 10.1121/1.4816541

11. Magee, M., Copland, D., and Vogel, A. P. (2019). Motor speech and non-motor language endophenotypes of Parkinson’s disease. Expert Rev. Neurother. 19, 1191–1200. doi: 10.1080/14737175.2019.1649142

12. Hlavnička, J., Čmejla, R., Tykalová, T., Šonka, K., Růžička, E., and Rusz, J. (2017). Automated analysis of connected speech reveals early biomarkers of Parkinson’s disease in patients with rapid eye movement sleep behaviour disorder. Sci. Rep. 7:12. doi: 10.1038/s41598-017-00047-5

13. Costantini, G., Cesarini, V., Di Leo, P., Amato, F., Suppa, A., Asci, F., et al. (2023). Artificial intelligence-based voice assessment of patients with Parkinson’s disease off and on treatment: machine vs. deep-learning comparison. Sensors 23:2293. doi: 10.3390/s23042293

14. Pah, N. D., Motin, M. A., and Kumar, D. K. (2022). Phonemes based detection of Parkinson’s disease for telehealth applications. Sci. Rep. 12:9687. doi: 10.1038/s41598-022-13865-z

15. Rahmatallah, Y., Kemp, A., Iyer, A., Pillai, L., Larson-Prior, L., Virmani, T., et al. (2024). Pre-trained convolutional neural networks identify Parkinson’s disease from spectrogram images of voice samples. Res. Sq. [Preprint]. Available at: doi:10.21203/rs.3.rs-5348708/v1 (Accessed: December 26, 2025).

16. Eguchi, K., Yaguchi, H., Kudo, I., Kimura, I., Nabekura, T., Kumagai, R., et al. (2024). Differentiation of speech in Parkinson’s disease and spinocerebellar degeneration using deep neural networks. J. Neurol. 271, 1004–1012. doi: 10.1007/s00415-023-12091-5

17. Mendes-Laureano, J., Gómez-García, J. A., Guerrero-López, A., Luque-Buzo, E., Arias-Londoño, J. D., Grandas-Pérez, F., et al. (2024). NeuroVoz: a Castillian Spanish corpus of Parkinsonian speech. Sci. Data 11:1367. doi: 10.1038/s41597-024-04186-z

18. Dimauro, G., Di Nicola, V., Bevilacqua, V., Caivano, D., and Girardi, F. (2017). Assessment of speech intelligibility in Parkinson’s disease using a speech-to-text system. IEEE Access 5, 22199–22208. doi: 10.1109/ACCESS.2017.2762475

19. Jadoul, Y., Thompson, B., and de Boer, B. (2018). Introducing Parselmouth: a Python interface to Praat. J. Phonetics 71, 1–15. doi: 10.1016/j.wocn.2018.07.001

20. Boersma, P., and Weenink, D. (2021). Praat: doing phonetics by computer (Version 6.1.38) [Computer program]. Available at: http://www.praat.org/ (Accessed: December 26, 2025).

21. Skodda, S., Visser, W., and Schlegel, U. (2011). Vowel articulation in Parkinson’s disease. J. Voice 25, 467–472. doi: 10.1016/j.jvoice.2010.01.009

22. He, K., Zhang, X., Ren, S., and Sun, J. (2016). "Deep residual learning for image recognition," in 2016 IEEE Conference on Computer Vision and Pattern Recognition (CVPR) (Las Vegas, NV: IEEE), 770–778. doi: 10.1109/CVPR.2016.90

23. Fisher, R. A. (1970). "Statistical methods for research workers," in Breakthroughs in Statistics: Methodology and Distribution (New York, NY: Springer), 66–70.

24. Selvaraju, R. R., Cogswell, M., Das, A., Vedantam, R., Parikh, D., and Batra, D. (2017). "Grad-CAM: visual explanations from deep networks via gradient-based localization," in 2017 IEEE International Conference on Computer Vision (ICCV) (Venice, Italy: IEEE), 618–626. doi: 10.1109/ICCV.2017.74.

25. Huzaifah, M. (2017). Comparison of time-frequency representations for environmental sound classification using convolutional neural networks. arXiv [Preprint]. Available at: https://arxiv.org/abs/1706.07156 (Accessed: December 26, 2025).

26. Hasegawa-Johnson, M., Pizza, S., Alwan, A., Cha, J. S., and Haker, K. (2003). Vowel category dependence of the relationship between palate height, tongue height, and oral area. J. Speech Lang. Hear. Res. 46, 738–753. doi: 10.1044/1092-4388(2003/059)

27. Mefferd, A. S., and Dietrich, M. S. (2019). Tongue- and jaw-specific articulatory underpinnings of reduced and enhanced acoustic vowel contrast in talkers with Parkinson’s disease. J. Speech Lang. Hear. Res. 62, 2118–2132. doi: 10.1044/2019_JSLHR-S-MSC18-18-0192

28. Sapir, S., Ramig, L., and Fox, C. (2008). Speech and swallowing disorders in Parkinson disease. Curr. Opin. Otolaryngol. Head Neck Surg. 16, 205–210. doi: 10.1097/MOO.0b013e3282febd3a

29. Wright, H., and Aharonson, V. (2025). Vocal feature changes for monitoring Parkinson’s disease progression—a systematic review. Brain Sci. 15:320. doi: 10.3390/brainsci15030320

